# Using the Mercy Method to Estimate Ideal Body Weight in Children – A Description and Validation of a New Methodology

**DOI:** 10.1101/2020.04.27.20081430

**Authors:** Mike Wells, Lara Nicole Goldstein

**Affiliations:** Division of Emergency Medicine, Faculty of Health Sciences, University of the Witwatersrand

## Abstract

**Background:** In obese children, lipophilic medications should be dosed to total body weight (TBW) and hydrophilic medications to ideal body weight (IBW). During emergencies these weights need to be estimated to ensure that urgent drug therapy is accurate and safe. The Mercy method is a well-established weight estimation system that has recently been adapted to provide estimations of body length in children. It was therefore conceivable that this could be further modified to provide estimations of IBW.

**Methods:** A model was developed *a priori* using the Mercy method’s humeral length segments to predict IBW. The accuracy of this model was then tested in a sample of 13134 children from the National Health and Nutrition Examination Survey (NHANES) datasets. The accuracy of IBW estimation was determined from the percentage of estimations falling within 10% (p10) and 20% (p20) of true IBW. The model was also tested to see the accuracy of detection of obesity in the study sample.

**Results:** From the sample of 13134 children, a subset of 1318 obese children were identified. In this subset, the new Mercy method model achieved an IBW estimation accuracy p10 of 66.9% and a p20 of 95.1%. For the detection of the obese child, the model had a sensitivity of 88.6% and a specificity of 75.8%.

**Conclusions:** This study established that the Mercy method can be modified to provide a reasonably accurate estimation of IBW in obese children, with very few critical errors. The ability to the model to identify the obese child was also reasonably accurate, on a par with other such predictive methods. While other accurate methods of estimating both TBW and IBW exist, such as the PAWPER XL tape, the modified Mercy method is an acceptable alternative if such other devices are not available.

## INTRODUCTION

### Background

The World Health Organisation has recommended that the doses of hydrophilic drugs be calculated according to ideal body weight (IBW) in obese children [1]. This is necessary in order to prevent potentially harmful overdoses of these medications in these children. A calculation or an estimation of IBW is therefore required for every obese child who requires drug treatment with hydrophilic drugs (e.g. adrenaline, electrolytes, sodium bicarbonate). Lipophilic drugs (e.g. amiodarone, benzodiazepines, anti-epileptic drugs), on the other hand, need to be calculated to total body weight (TBW) in obese children [1]. Therefore, both TBW and IBW need to be known in order to calculate drug doses safely in obese children [2].

During medical emergencies in children – when a child cannot be weighed to determine TBW and there is no time to calculate IBW – it is necessary to be able to estimate both these dose scalars before drugs can be administered [3]. Devices such as the PAWPER XL tape are able to estimate both TBW and IBW accurately in obese children, but the tape might not be available in all settings [4]. The Mercy method is another system which can estimate TBW accurately, with similar accuracy to the PAWPER XL tape, but it can be used with a standard tape measure [5, 6]. The Mercy method, however, currently has no mechanism with which to provide an estimation of IBW. Recent work showing how the Mercy method can be used to predict a child’s height, has provided a potential approach by which the system could also be used to estimate IBW [7].

### Importance

The ability to estimate IBW is important since many drugs used in the resuscitation of critically ill or injured children are hydrophilic, and since poor outcomes in obese children have been ascribed to dosing errors based on TBW estimations [8, 9]. An additional method that could be used to estimate both TBW and IBW would therefore be useful for clinicians caring for sick children requiring emergency care.

### Objectives

The objectives of this study were to develop a model by which the Mercy method could be adapted to provide an estimation of IBW; to validate this model in a large database; and to develop tables that could be used by clinicians to estimate TBW as well as IBW by using the standard Mercy measurements.

## METHODS

### Description of the Mercy method

The Mercy method, which is validated in children up to the age of 16 years, makes use of two anthropometric measurements to generate an estimate of TBW [10]. Measurements of humeral length (HL) and mid-arm circumference (MAC) are obtained and a “partial weight” read off a table for each measurement (Figure 1). The predicted TBW is then obtained from the sum of these two “partial weights”.

**Figure 1:**
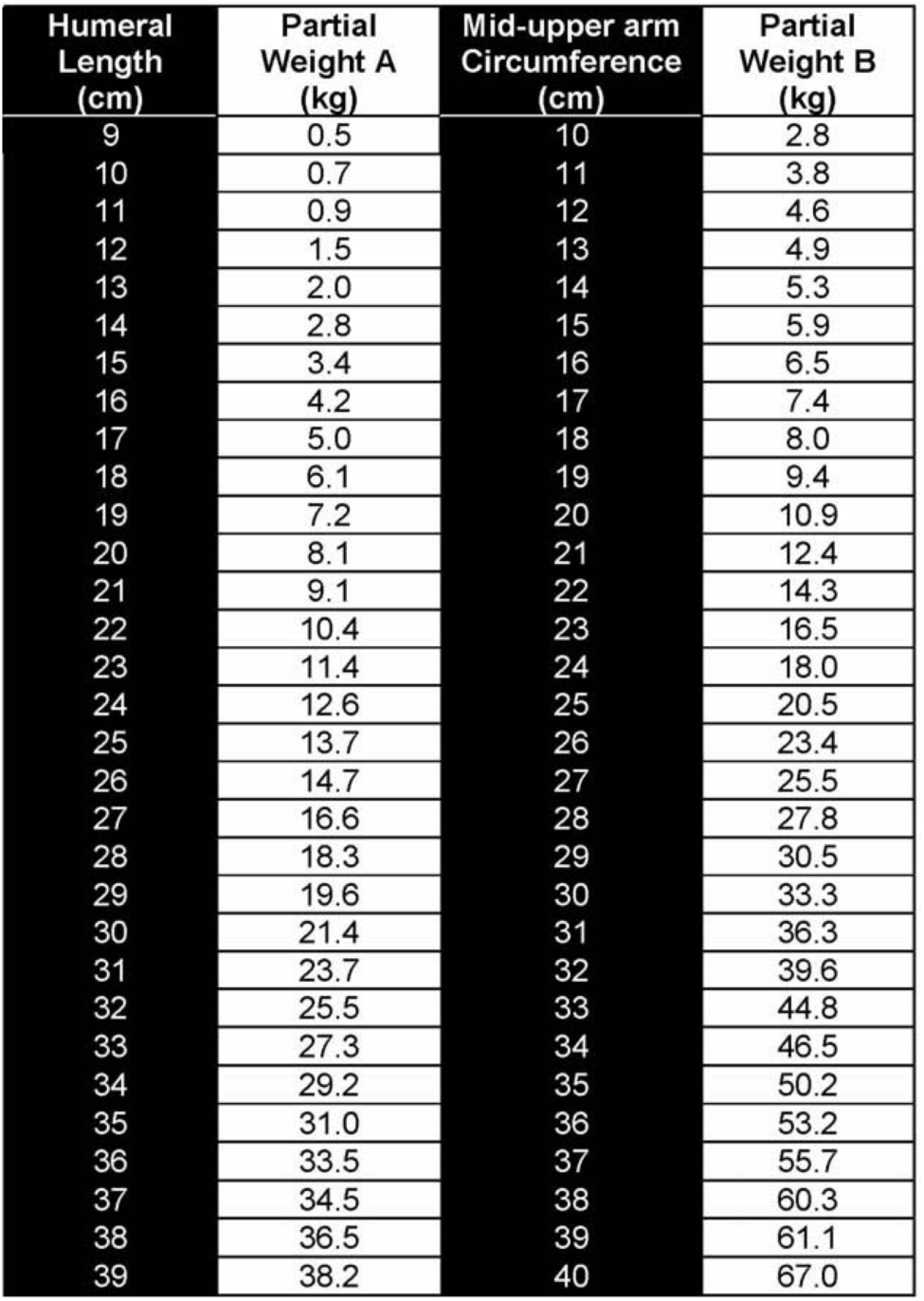
The Mercy method chart of partial weights. This is the figure that was provided in the first published description of the Mercy method (CC by 3.0 Abdel-Rahman and Ridge) [10]. Once the user has obtained measurements of humeral length and mid-arm circumference (or mid-upper arm circumference) then the partial weights can be read off and added together to obtain the estimate of total body weight (kg).

### Development of the model to predict IBW

The publication of a validated method to estimate children’s length from humeral length created the possibility of predicting IBW from humeral length [7]. Our model to predict IBW from measurements of humeral length was developed as follows:

- Firstly, the recumbent lengths predicted by the Mercy system for each centimetre increment of humeral length were collated into an electronic worksheet (Microsoft Excel for Mac Version 16.35 (2020), Microsoft, Redmond, Washington) [7].
- Secondly, since IBW is very closely related to recumbent length, it was possible to create a table of IBWs for each range of humeral lengths [11]. This was done by using published data available from the PAWPER XL-MAC tape system to calculate the IBW for each humeral length category [12]. The PAWPER XL system has been shown to be able to predict IBW within 10% in 100% of obese children [2].
- Thirdly, the IBW value in each category was cross-checked against an IBW calculated from length using the Traub-Kichen formula [13]. If the difference between this and the PAWPER XL-MAC value was greater than 5%, then an average was used for the IBW.
- Fourthly, since IBW should only be used in obese children, a threshold value for TBW was calculated to inform the user which child would be classified as obese, in which case the value for IBW would be relevant. This threshold value was calculated in two ways. The PAWPER XL-MAC weight for a child with a habitus score of 5 (obese) was identified for each length-group from previously published data as an indicator of an obese weight [12]. In addition, a weight of 120% of the IBW for each length-category was calculated – this is a value which has been reported to be predictive of obesity [14]. If the difference between this and the PAWPER XL-MAC value was greater than 5%, then an average was used for the threshold.
- Finally, spreadsheet formulas were created with the Mercy method humeral length categories, the predicted recumbent length, the predicted IBW, and the predicted TBW at which the child should be considered to be obese.

These formulas were then used to calculate the output data for the validation component of the study.

### Validation of the IBW model

#### Datasets

The National Health and Nutrition Examination Survey (NHANES) datasets A, B, C, D and G (datasets from the 1999–2000 to 2011–2012 surveys) were obtained from the Centers for Disease Control (CDC) website [15]. The demographic and anthropometric datasets were merged and all data for children ≤16 years of age were extracted. The extracted variables included age, sex, height or recumbent length, total body weight, BMI, MAC and humeral length. Cases with missing data were excluded.

BMI-for-age Z-scores were calculated for each child using World Health Organisation (WHO) reference data for children under the age of 24 months and CDC reference data for children from 2 to 16 years of age. Data for children with a BMI-for-age Z-score of ≥2.0 (obese children) were extracted for further analysis.

The IBW_50_ method was used as the reference method for IBW in this study and was calculated as the weight at the 50^th^ centile of BMI-for-age for each child. This method is generally regarded as the most accurate and useful of the methods of determining IBW [11].

#### Weight estimation generation

Estimates of TBW were generated using the standard methodology of the Mercy method from the measurements of humeral length and MAC. Estimates of IBW were generated using the model developed in this study from the measurements of humeral length.

### Data analysis

The success of IBW estimation of the model was evaluated using statistical methods centred on percentage error analysis. Three primary statistical measures were used to assess performance: mean percentage error (MPE) indicated the estimation bias; the 95% limits of agreement of the MPE (PELOA) represented the estimation precision; and the percentage of weight estimations that fell within 10% (PW10) and 20% (PW20) of calculated IBW_50_ denoted overall accuracy.

The performance of TBW estimation of the Mercy method was also evaluated using the same methodology.

Subgroup analyses of the performance of the Mercy method were performed in age-categories: ≤2 years, 2.1 to 5 years, 5.1 to 10 years and >10 years.

The accuracy of the identification of obese children by the model was evaluated by determining the sensitivity and specificity of the model (performed in the entire pooled dataset). A a BMI-for-age Z-score threshold of 2.0 was used as a definition of obesity.

The primary outcome measure was the performance of the model when compared to calculated IBW_50_. A PW10 >70% and a PW20 >95% was considered to be an acceptable accuracy of estimation [5]. This benchmark is the level of accuracy regularly achieved by the dual length-based, habitus-modified weight-estimation systems.

### Creation of the tables for the validated model

Following the validation of the model, a set of tables was created that could be used by clinicians to estimate TBW and IBW using the modified Mercy method.

## RESULTS

### Demographic data

There were 13134 children in the pooled database. Of these children 1318 met the criteria for the weight estimation accuracy component of the study (age ≤16 years, BMI-for-age ≥2.0, complete data). There were 616 males (46.7%) and 702 females (53.3%). The rest of the demographic data is summarised in Table 1.

**Table 1:**
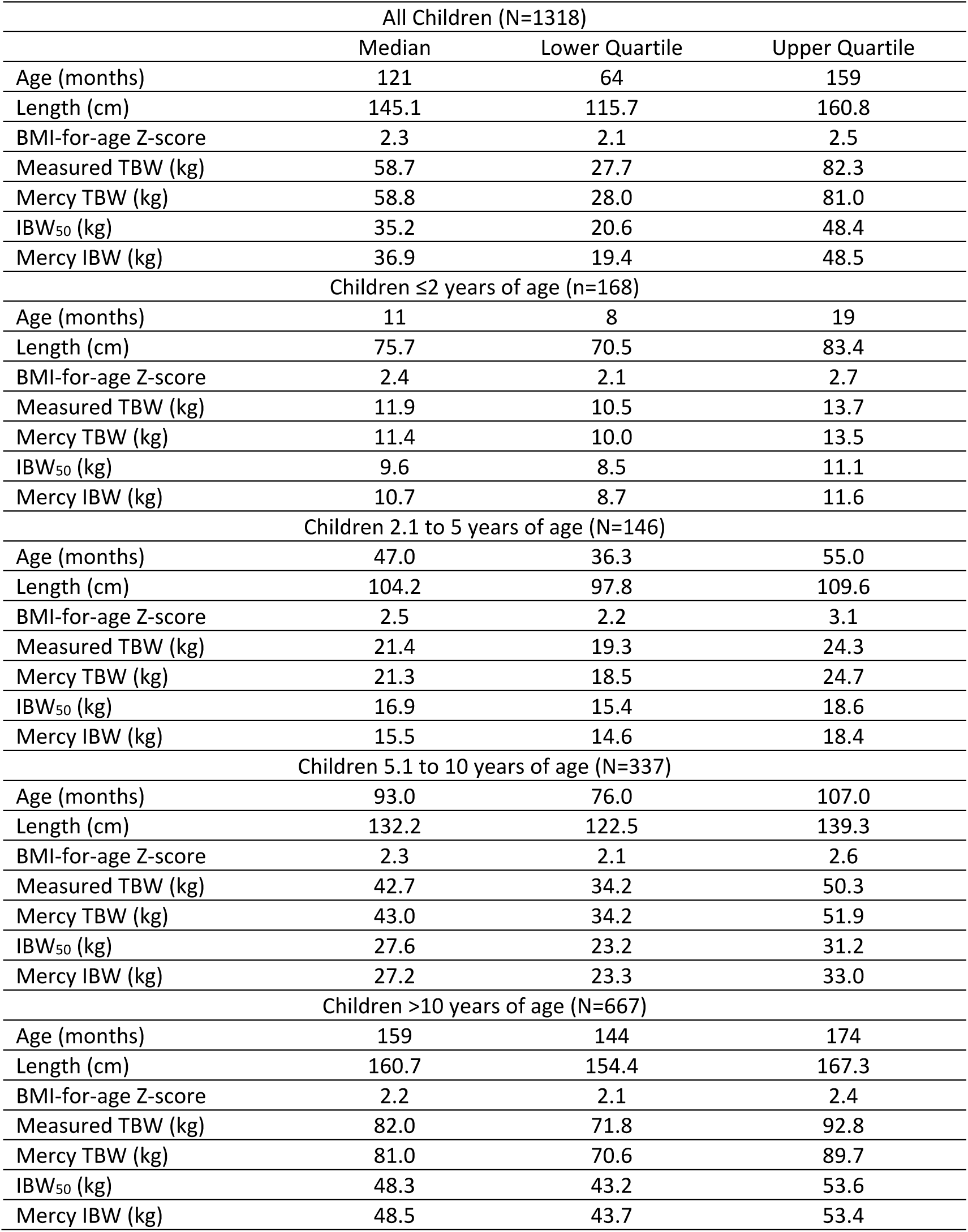
Demographic and anthropometric data for the study population. The data is shown for the whole sample as well as by subgroup of age. Medians with upper and lower quartiles are displayed. Abbreviations: BMI - body mass index; TBW - total body weight; IBW_50_ - standard calculated ideal body weight using the BMI method.

| All Children (N=1318) |  |  |  |
| --- | --- | --- | --- |
|  | Median | Lower Quartile | Upper Quartile |
| Age (months) | 121 | 64 | 159 |
| Length (cm) | 145.1 | 115.7 | 160.8 |
| BMI-for-age Z-score | 2.3 | 2.1 | 2.5 |
| Measured TBW (kg) | 58.7 | 27.7 | 82.3 |
| Mercy TBW (kg) | 58.8 | 28.0 | 81.0 |
| IBW<sub>50</sub> (kg) | 35.2 | 20.6 | 48.4 |
| Mercy IBW (kg) | 36.9 | 19.4 | 48.5 |
| Children ≤2 years of age (n=168) |  |  |  |
| Age (months) | 11 | 8 | 19 |
| Length (cm) | 75.7 | 70.5 | 83.4 |
| BMI-for-age Z-score | 2.4 | 2.1 | 2.7 |
| Measured TBW (kg) | 11.9 | 10.5 | 13.7 |
| Mercy TBW (kg) | 11.4 | 10.0 | 13.5 |
| IBW<sub>50</sub> (kg) | 9.6 | 8.5 | 11.1 |
| Mercy IBW (kg) | 10.7 | 8.7 | 11.6 |
| Children 2.1 to 5 years of age (N=146) |  |  |  |
| Age (months) | 47.0 | 36.3 | 55.0 |
| Length (cm) | 104.2 | 97.8 | 109.6 |
| BMI-for-age Z-score | 2.5 | 2.2 | 3.1 |
| Measured TBW (kg) | 21.4 | 19.3 | 24.3 |
| Mercy TBW (kg) | 21.3 | 18.5 | 24.7 |
| IBW<sub>50</sub> (kg) | 16.9 | 15.4 | 18.6 |
| Mercy IBW (kg) | 15.5 | 14.6 | 18.4 |
| Children 5.1 to 10 years of age (N=337) |  |  |  |
| Age (months) | 93.0 | 76.0 | 107.0 |
| Length (cm) | 132.2 | 122.5 | 139.3 |
| BMI-for-age Z-score | 2.3 | 2.1 | 2.6 |
| Measured TBW (kg) | 42.7 | 34.2 | 50.3 |
| Mercy TBW (kg) | 43.0 | 34.2 | 51.9 |
| IBW<sub>50</sub> (kg) | 27.6 | 23.2 | 31.2 |
| Mercy IBW (kg) | 27.2 | 23.3 | 33.0 |
| Children >10 years of age (N=667) |  |  |  |
| Age (months) | 159 | 144 | 174 |
| Length (cm) | 160.7 | 154.4 | 167.3 |
| BMI-for-age Z-score | 2.2 | 2.1 | 2.4 |
| Measured TBW (kg) | 82.0 | 71.8 | 92.8 |
| Mercy TBW (kg) | 81.0 | 70.6 | 89.7 |
| IBW<sub>50</sub> (kg) | 48.3 | 43.2 | 53.6 |
| Mercy IBW (kg) | 48.5 | 43.7 | 53.4 |

### TBW and IBW performance data

The bias, precision and accuracy data of the Mercy method in predicting TBW and IBW in this study population is shown in Table 2 for the population as a whole and for each of the subgroups.

**Table 2:** Results of the analysis of the performance of the Mercy method. The table shows the performance of the Mercy method in estimating total body weight and the modified Mercy method in estimating ideal body weight. Abbreviations: MPE - mean percentage error (a measure of bias); LLOA - lower limit of agreement; ULOA - upper limit of agreement (the 95% limits of agreement are a measure of precision); p10 - percentage of estimates falling within 10% of the index weight (measured weight and calculated ideal body weight); p20 - percentage of estimates falling within 20% of the index weight (these are measures of overall accuracy).

|  | N | TOTAL BODY WEIGHT ESTIMATIONS |  |  |  |  | IDEAL BODY WEIGHT ESTIMATIONS |  |  |  |  |
| --- | --- | --- | --- | --- | --- | --- | --- | --- | --- | --- | --- |
|  |  | MPE (%) | LLOA (%) | ULOA (%) | p10 (%) | p20 (%) | MPE (%) | LLOA (%) | ULOA (%) | p10 (%) | p20 (%) |
| All Children | 1318 | -1.3 | -20.6 | 18.0 | 73.8 | 96.1 | 0.0 | -20.8 | 20.9 | 66.9 | 95.1 |
| Children ≤2 years | 168 | -3.1 | -26.6 | 20.4 | 60.1 | 91.7 | 2.5 | -21.2 | 26.1 | 56.0 | 91.7 |
| Children 2.1 to 5 years | 146 | -1.7 | -19.3 | 15.9 | 75.3 | 97.9 | -0.7 | -19.1 | 17.7 | 73.3 | 96.6 |
| Children 5.1 to 10 years | 337 | 1.5 | -18.1 | 21.1 | 76.3 | 96.1 | 2.3 | -18.7 | 23.2 | 69.4 | 95.3 |
| Children >10 years | 667 | -2.2 | -20.0 | 15.5 | 75.7 | 96.9 | -1.6 | -21.6 | 18.5 | 67.0 | 95.7 |

#### Obesity identification accuracy data

The sensitivity and specificity of the model used to predict obesity (a TBW >120% of IBW) is shown in Table 3.

**Table 3:** Sensitivity and specificity of the obesity threshold weight. This analysis was performed in the entire sample of children from the dataset (obese and non-obese) to evaluate the sensitivity and specificity of the model. A threshold of 120% of ideal body weight was selected - if a child’s total body weight exceeded this value then the child was defined as obese.

|  | **N** | **Sensitivity (%)** | **Specificity (%)** |
| --- | --- | --- | --- |
| **All Children** | 13132 | 88.6 | 75.8 |
| **Children $\leq 2$ years of age** | 1686 | 31.4 | 92.9 |
| **Children 2.1 to 5 years of age** | 2081 | 63.5 | 93.9 |
| **Children 5.1 to 10 years of age** | 2602 | 94.9 | 83.1 |
| **Children $>10$ years of age** | 6762 | 100 | 66.9 |

### Chart for clinical use

The table with the predicted IBW for each humeral length, as well as the cut-off values for TBW at which IBW should be used, are shown in Figure 2.

**Figure 2:**
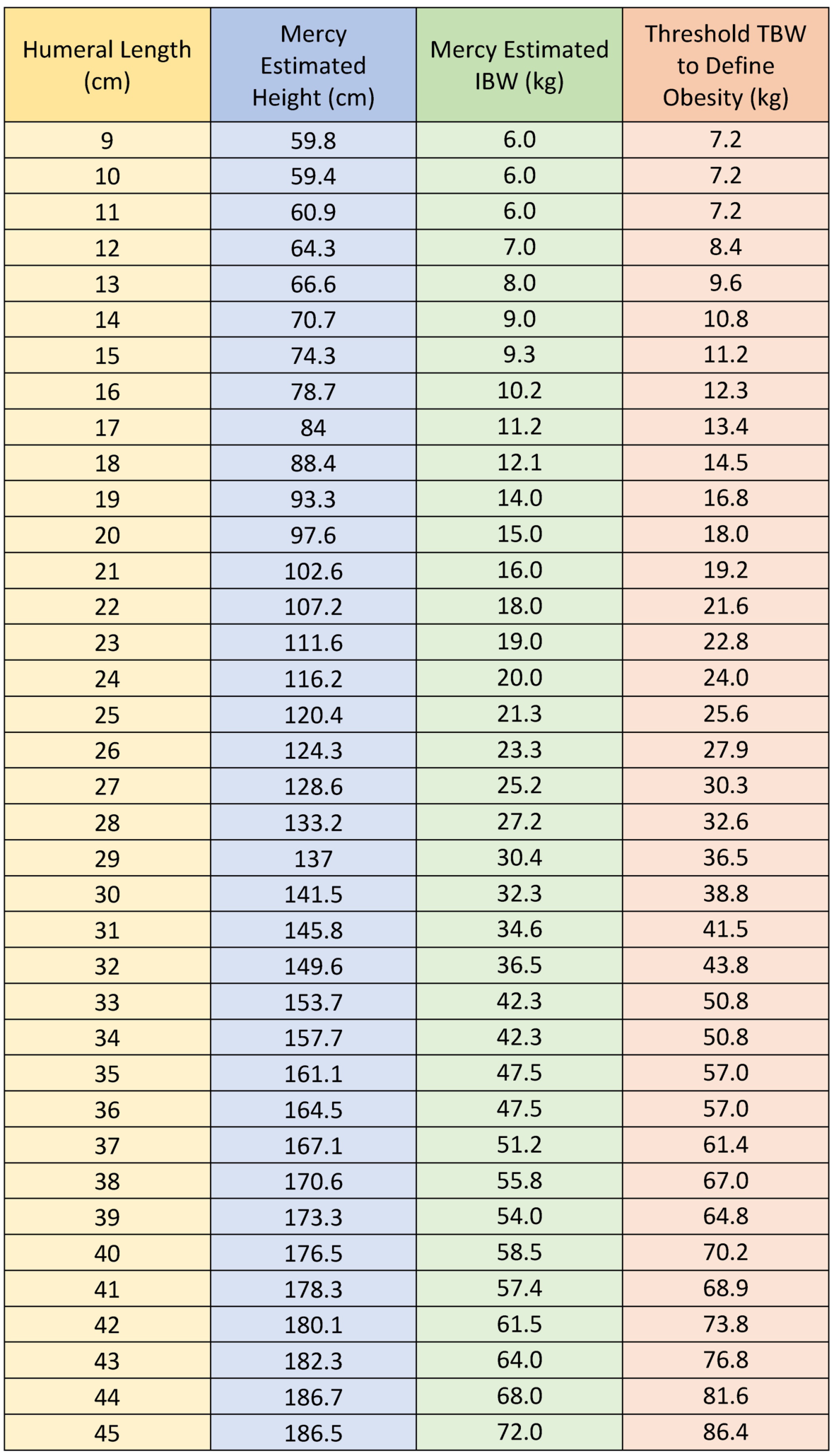
Ideal body weights and threshold weights to define obesity for each humeral length category. After obtaining a measurement of a child’s humeral length and mid-arm circumference, the Mercy method total body weight can be determined using the standard methodology. This table can then be used to identify the obese child: if the total body weight exceeds the threshold weight at the child’s humeral length then that child is obese. The ideal body weight corresponding to the correct humeral length can be read off the table. The ideal body weight would be used for hydrophilic drug dose calculations while the total body weight would still be used for lipophilic drug dose calculations.

## DISCUSSION

Ideally a weight estimation system should be able to provide an accurate estimation of both TBW and IBW in obese children to allow for appropriate and accurate drug dose calculations [12]. The Mercy method has already been shown to be accurate in providing estimates of TBW in children in both high-income and low- and middle-income settings [5, 16]. The Mercy method proved in this current study, that its methodology can be adapted to predict IBW with a reasonable degree of accuracy. This methodology also showed promise in identifying obese children, for whom this methodology would be used.

There are several other systems that have been evaluated for emergency estimation of IBW in obese children, with which to compare the performance of the novel Mercy method. Age-based formulas, despite speculation by some authors that they might be accurate, have proven to be poor at estimating IBW in obese children (unpublished data: Wells M, Goldstein L: The utility of pediatric age-based weight estimation formulas for emergency drug dose calculations in obese children. Journal of the American College of Emergency Physicians Open. 2020, In Press.). These formulas could not achieve a p10 of more than 50% (compared to the modified Mercy method in this study of just below 70%). Age formulas also had a high critical error rate of around 20%, compared to the 5% of the modified Mercy method. Given the poor accuracy of age formulas in predicting TBW and IBW, they should not be used [17]. The Broselow tape and the PAWPER XL tape have both been evaluated in terms of their ability to predict IBW [3]. They are length-based and length- and habitus-based weight estimation systems and have achieved extremely accurate estimations of IBW, far superior to the Mercy method. The Broselow tape, however, has a poor accuracy for estimating TBW and is restricted to children under 12 years of age [18]. The PAWPER XL tape, on the other hand, is able to estimate both TBW and IBW very accurately: estimations of IBW are far superior to that achieved by the Mercy method, and the estimations of TBW are similar to or slightly better than those of the Mercy method [5]. The Mercy method could, therefore, justifiably be used as an alternative to the PAWPER XL tape for both TBW and IBW estimation if the tape was not available.

The ability of this model to determine when IBW should be used (i.e. to predict obese children) was good in children over the age of two years, and very good in children over the age of five years. This aspect (the accuracy of identification of obesity) has not been evaluated in any previous weight estimation system. The sensitivity and specificity of the model is similar to that in other models used to predict obesity in body composition studies [19]. Future fine tuning of the model could also potentially increase its discriminatory ability. The use of a system such as this will become more important in the future as clinicians focus on calculating the right dose for each drug for each child, depending on their body habitus. The increasing prevalence of childhood obesity in both high-income as well as low- and middle-income countries require that emergency medicine doctors have the knowledge and resources to optimise drug dosing in obese children. This is an important matter of patient safety.

There are some potential limitations to using the Mercy method in emergency care. Firstly, the correct use of the Mercy method – and therefore its accuracy – is highly dependent on appropriate training and experience of the users [20]. Secondly, the accuracy of Mercy method has been shown to be vulnerable when used during simulated emergencies as the measurement techniques might not be optimal as a result of the positioning and cooperation of the child and the stressful environment [21, 22]. Finally, the Mercy method has been shown previously to underperform in infants and young children and should be used with care in this group of patients [23].

### Limitations

This study was performed in part of the dataset that was used for the development of the Mercy method originally. For this reason, the TBW estimates may be slightly more accurate than may be expected in other study samples. This should have a much smaller impact on the IBW model developed in the current study. In addition, although virtual studies performed in databases have been shown to be valid, the accuracy of the IBW predictions will need to be established in future prospective studies [24].

## CONCLUSIONS

These findings advance the understanding of the methods of estimation of IBW in children when it cannot be calculated electively. This study established that the Mercy method can be modified to provide a reasonably accurate estimation of IBW in obese children, with very few critical errors. This methodology was substantially more accurate than age-based formulas but was markedly less accurate than the Broselow tape and the PAWPER XL tape, according to data from previous studies. Since the Mercy method is able to predict TBW significantly more accurately than age-formulas and the Broselow tape, its use would be recommended over these methodologies.

The performance of the Mercy method and the PAWPER XL tape are similar with respect to TBW, according to previous studies. Although the PAWPER XL tape is significantly more accurate at predicting IBW than the Mercy method, both have a low enough critical error rate that they could be used confidently in clinical practice after further validation.

The ability to the model to accurately identify the obese child was reasonable, but additional work in the future would be required to refine this methodology. This was the first study to evaluate the ability of a weight estimation system to predict in which children the use of IBW should be considered, for hydrophilic drug dose calculations.

## Data Availability

Datasets are open access.

https://www.cdc.gov/nchs/nhanes/continuousnhanes/default.aspx

